# “I didn’t even know humans could get polio … I swear that’s for dogs.” A co-produced, thematic analysis exploring adolescent attitudes to vaccines

**DOI:** 10.1101/2025.10.29.25338465

**Authors:** Angie Pitt, Richard Amlôt, Catherine Heffernan, G. James Rubin, Louise E. Smith

## Abstract

Immunisation forms a cornerstone of public health policy. However, uptake rates of routine vaccines offered in adolescence are sub-optimal, presenting both individual and public health risks.

We worked with over 80 adolescents to co-design, pilot and analyse data for this qualitative analysis of adolescent attitudes. For the study itself we interviewed 30 adolescents aged 12-17 years and living in England. Five themes were identified: Understanding vaccines, Fear and comfort; Autonomy and control; Information sources and conspiracy theories; and Identity and Social norms.

Among our participants, vaccine uptake was influenced by perceptions of vaccine preventable diseases and by fear of needles. Decision-making dynamics between parents and adolescents were complex, with autonomy disrupting as well as driving vaccine behaviour. Social media was not a major source of vaccine information for adolescents, with participants expressing disinterest in online vaccine content and implicating adults as the ones engaging with and sharing online vaccine disinformation. Finally, we found that adolescent development impacted vaccine attitudes. Younger participants were more likely to adopt a collective family attitude toward vaccines. Older participants were more able to consider long-term benefits and more likely to have established their own position on vaccines.

Our findings highlight the need for adolescents to be: involved in vaccine decisions; given techniques to manage anxiety about vaccines; better educated about vaccine preventable diseases; and supported in developing the critical thinking skills needed to make informed health decisions.

## Introduction

Immunisation forms a cornerstone of UK public health policy, with adolescence being a key stage in the Routine Immunisation Schedule [1]. Between the ages of 11- and 16-years adolescents in England are offered several vaccines: MenACWY (for meningitis), Td/IPV (for tetanus, diphtheria and polio, HPV (human papilloma virus which protects against some cancers and genital warts) alongside seasonal influenza vaccines. Routine vaccines are offered in school, with primary healthcare providers offering a catch-up service for those who miss school vaccines. From 2021, a Covid-19 vaccine was offered to UK adolescents aged 12 to 17 in response to the pandemic; however, this age group were among the most hesitant to receive it [2]. Covid-19 vaccines are now offered only to adolescents with weakened immune systems [4]. Uptake of routine adolescent vaccines, which had already been decreasing year on year since 2012 [6], dropped further during the COVID-19 pandemic [6]. In 2023/24, HPV uptake in England, for example, was 72.9% of girls and 67.7% of boys in school year 8 (aged 12 to 13 years), well below the World Health Organization’s goal of achieving 90% coverage by 2040 [7]. Consequently, in 2025 NHS England launched a catch-up campaign targeting HPV-unvaccinated individuals aged 16-24 [8]. While much research exists on the attitudes of healthcare workers [9] and parents [10, 11], less is heard from adolescents themselves [3, 12].

Several psychosocial factors have been linked to adolescent vaccine behaviour. Fear may be significant, with fear of needles [13] and both parental [11, 14] and adolescent [15] concern about side effects implicated in hesitancy. In adults, vaccination side effects are more likely to be experienced by those expecting adverse effects, or observing adverse effects in others [16]. However, little research has explored how adolescents view side effects, and whether social influences impact side effect experiences.

Adolescence is also a period of intense developmental change and social redefinition [17, 18]. Adolescent decision-making is subject to social influence from peers [19] and a complex web of information sources including parents, school, and social media [3]. Previous literature has called for a better understanding of the role of information sources in adolescent vaccine decision-making [3, 20].

Knowledge and autonomy have been linked to vaccine uptake [21, 22]. Evidence suggests increasing vaccine knowledge improves adolescent attitudes toward vaccines [23] although knowledge alone does not appear to drive uptake [24]. Knowledge is also linked with vaccine decision-making, with greater knowledge leading to increased involvement in vaccine decision-making [23, 25], and involvement in decision-making has also been linked to vaccine acceptance [25]. In England, notifications of upcoming vaccines are typically sent to parents or legal guardians, who are required to consent on behalf of their child. Adolescents aged 16 years and older can legally consent to vaccination, and adolescents <16 years can self-consent to vaccination if deemed cognitively competent (Gillick competent). However, in practice Gillick competence is rarely implemented with the role of parents as decision-makers prioritised over adolescent autonomy [26].

Health behaviours established in adolescence play a role in adulthood [27]. Consequently, adolescent vaccine hesitancy has the potential to negatively impact not only individual health, but also long-term public health in the UK. This study focuses on understanding views of 12– 17-year-old adolescents as the age range who have most recently been offered vaccinations. It explores how they view side effects, their sources of information and what influences their decision making so that we better address them in current and future public health campaigns.

### Methodology, incorporating Patient and Public Involvement and Engagement (PPIE)

This study was approved by the King’s College London Research Ethics Committee (reference LRS/DP-23/24-39491). We have used the term ‘parents’ in this study to describe caregivers and legal guardians. We involved adolescents at design, piloting and analysis stages of this study, working in partnership with a PPIE team of 16- and 17-year-old psychology and sociology A-level pupils, who led piloting with 12- to 15-year-olds. In total over 80 adolescents from two local secondary schools in London worked with us on this study. Depending on their level of involvement, adolescents were thanked for PPIE-input with vouchers and school merits, plus in-kind benefits including work experience, careers advice and letters of acknowledgement. Table 1 outlines PPIE team-led input into the study.

**Table 1:**
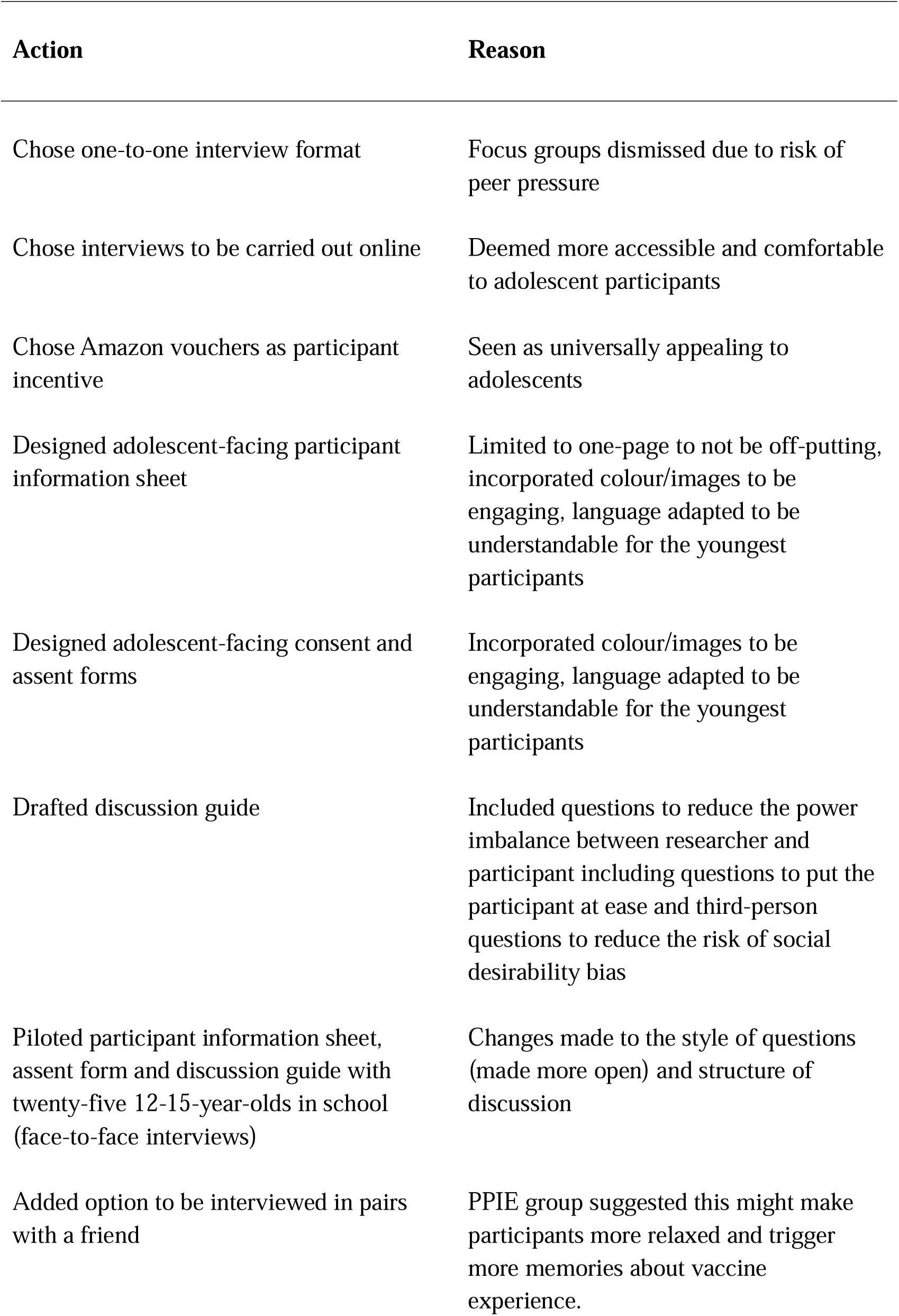
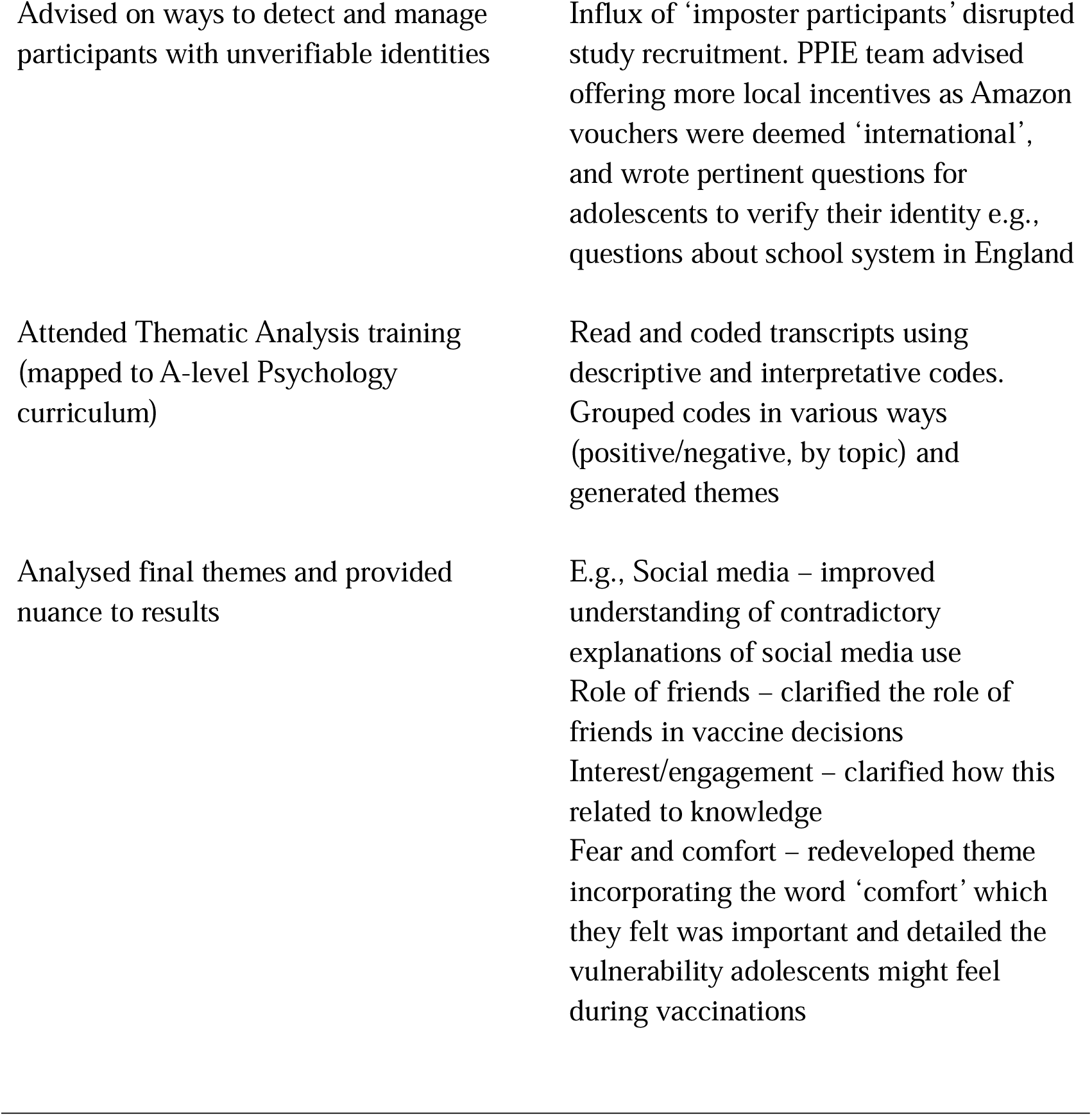
Patient and Public Involvement and Engagement (PPIE): Decisions & changes

### Design

Together we chose a qualitative design to capture the nuance, contradictions and rich detail of adolescent attitudes. The PPIE team chose structured interviews, rejecting focus groups over concerns about peer pressure.

### Participants and recruitment

Adolescents aged between 12- and 17-years (either in school or electively home educated) and living in England were eligible to take part. We aimed to interview 30 participants based on the principle of *information power* [28], which argues that the aim of the study, use of established theory, quality of dialogue and analysis strategy impact sample size. While our discussion was mapped to psychological theory, we anticipated a heterogenous sample given differences in adolescent development, family settings, and education. In line with Braun and Clarke [29], the concept of data saturation was not applied to this study. Braun & Clarke argue that in reflexive Thematic Analysis (TA) the researcher is responsive in interviews rather than following a rigid structure, that theme generation is iterative and not necessarily linear and new insights can be made while data continues to be collected.

The PPIE team designed a participant information sheet, consent and assent forms. We recruited participants using snowball and purposeful sampling via emails to schools and youth organisations, social media adverts, and via word of mouth. After challenges recruiting vaccine hesitant adolescents, we worked with a market research agency to recruit six additional participants. All participants completed an online screening survey via Qualtrics and completed consent/assent forms. Participants aged 16-17 years gave informed written consent. Where participants were aged 12-15 years, parents gave informed written consent and participants gave informed written assent. All participants also consented/assented verbally at the start of the interview.

### Discussion guide development

The PPIE team and researchers wrote the discussion guide together. The PPIE team felt social desirability bias, that is where participants give what they believe to be the expected answers [30], was a particular risk in research with adolescents. To mitigate this, and achieve a more balanced power dynamic, we included an emphasis that there were no right or wrong answers, that adolescents were the experts on their own experiences, and assuring confidentiality. Further, we included third person questions, e.g., “why might teenagers in general refuse vaccines?” to elicit non-biased answers. The PPIE team then planned and delivered a pilot day in their school to test, adapt and retest the information sheets, assent forms and discussion guide with pupils aged 12-15 years. No data were recorded during piloting. Finally, the research team mapped the discussion guide to the COM-B framework [31] to structure the discussion and ensure all potential psychological factors were explored. COM-B proposes behaviour is driven by capability, opportunity and motivation, and was chosen as it allows consideration of external influences on behaviour as well as individual motivations.

### Screening and data collection

Researchers communicated with participants via email, including parents in emails where participants were <16 for safeguarding. Following an influx of people attempting to participate with unverifiable or suspicious identities, the PPIE team advised on methods to detect potentially suspicious participants (Table 1). AP, a female PhD student with substantial experience working with this age group, interviewed all participants using Microsoft Teams. Participants were asked to switch on cameras temporarily to verify their identity and then could switch off their cameras if they wanted to. After piloting in-person interviews, online interviews were chosen for data collection as they were deemed safer, more accessible and less daunting for adolescents, as well as easier for adolescents to fit in alongside school and after-school clubs [32]. Interviews were video-recorded and transcribed by a third-party company. We asked participants to take part without a parent to ensure their privacy. AP made notes during and after interviews and used a journal to reflect on interviews. Participants were given a £20 Amazon voucher as a thanks for their time. Data were collected between November 2023 and March 2025.

### Data analysis

Data were analysed using reflexive thematic analysis [33] which allows an opportunity to locate participants’ experiences within wider socio-cultural [34]. We adopted a critical realist approach to understand not just what adolescents think about vaccines, but also how these attitudes develop alongside intense physical, neurological and social changes in adolescence [35]. We explored reflexivity through a journal and group discussion with the PPIE team, acknowledging our own biases toward vaccines. Data were analysed by both vaccine hesitant and vaccine acceptant PPIE team members and researchers. In line with Braun and Clarke [33] we continuously developed insights during data collection. In 2024, we trained the PPIE team in thematic analysis. Working together, we familiarised ourselves with the data through repeated readings, followed by line-by-line coding of the first 20 transcripts, recording both descriptive and interpretative codes relevant to the research questions. Across several sessions, the PPIE team organised codes into different groups, and eventually developed themes. Codes were added to NVivo [36] and the remaining transcripts were coded by AP according to the PPIE-co-developed codebook. Final themes were then taken back to the PPIE team for discussion and consensus. AP wrote the first draft of this report following the Consolidated Criteria for Reporting Qualitative Research (COREQ) guidelines [37].

## Results

### Participant characteristics

Thirty adolescent participants aged 12-17 years (Mdn = 13) took part in this study. Nineteen identified as female, the remainder identified as male (n = 10) or non-binary (n = 1). The majority were white (n = 21) and from London or the Southeast of England (n = 18). Ten participants had refused at least one secondary school-age routine vaccination so far, and a further five had refused some or all Covid vaccines (Table 2). The remainder had accepted all vaccines offered to them so far in adolescence. Interviews lasted an average of 36 minutes (range = 24 to 52 minutes). One participant had a parent present in the room (but not on camera) due to anxiety; the remaining participants were interviewed without a parent.

**Table 2:**
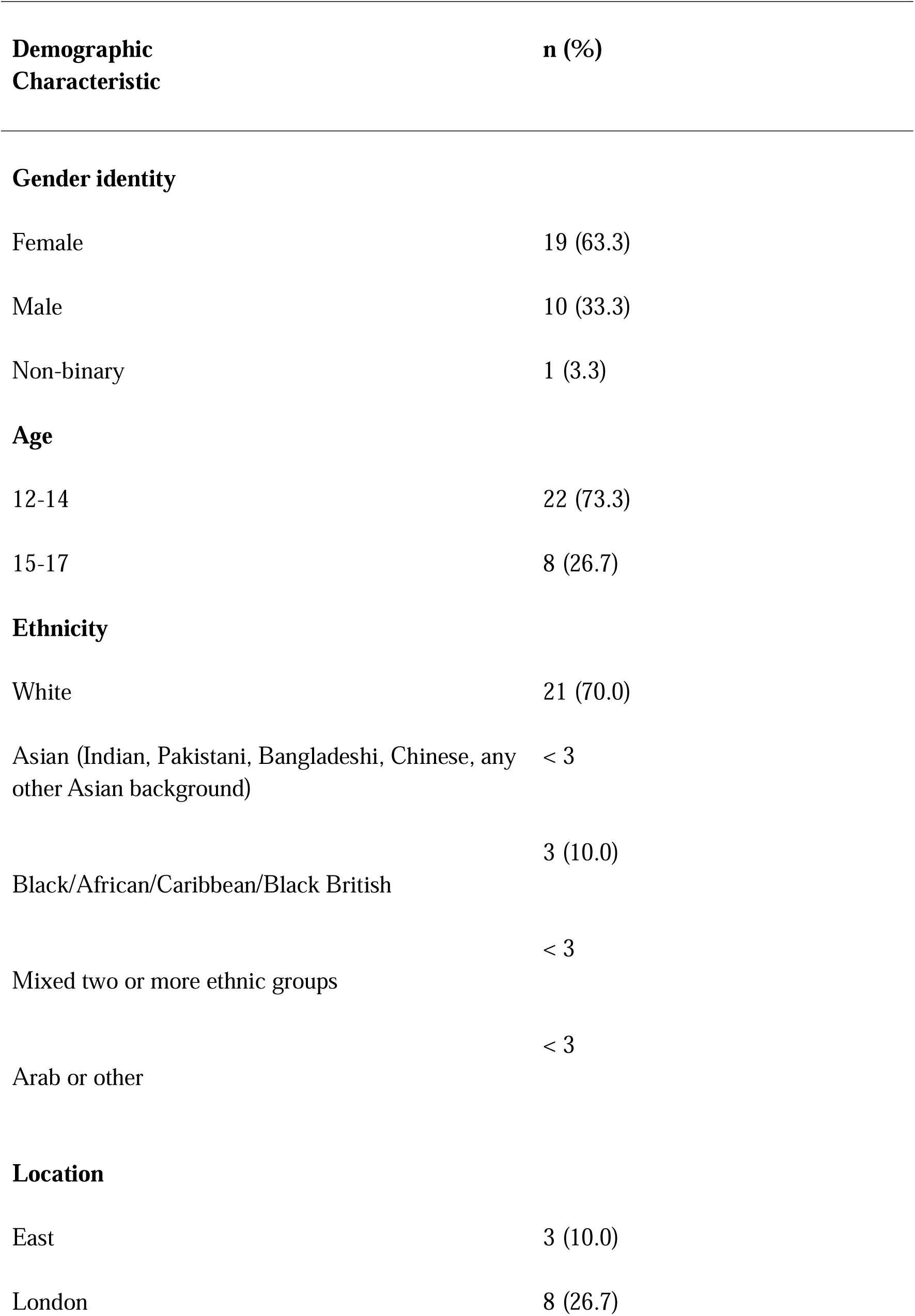
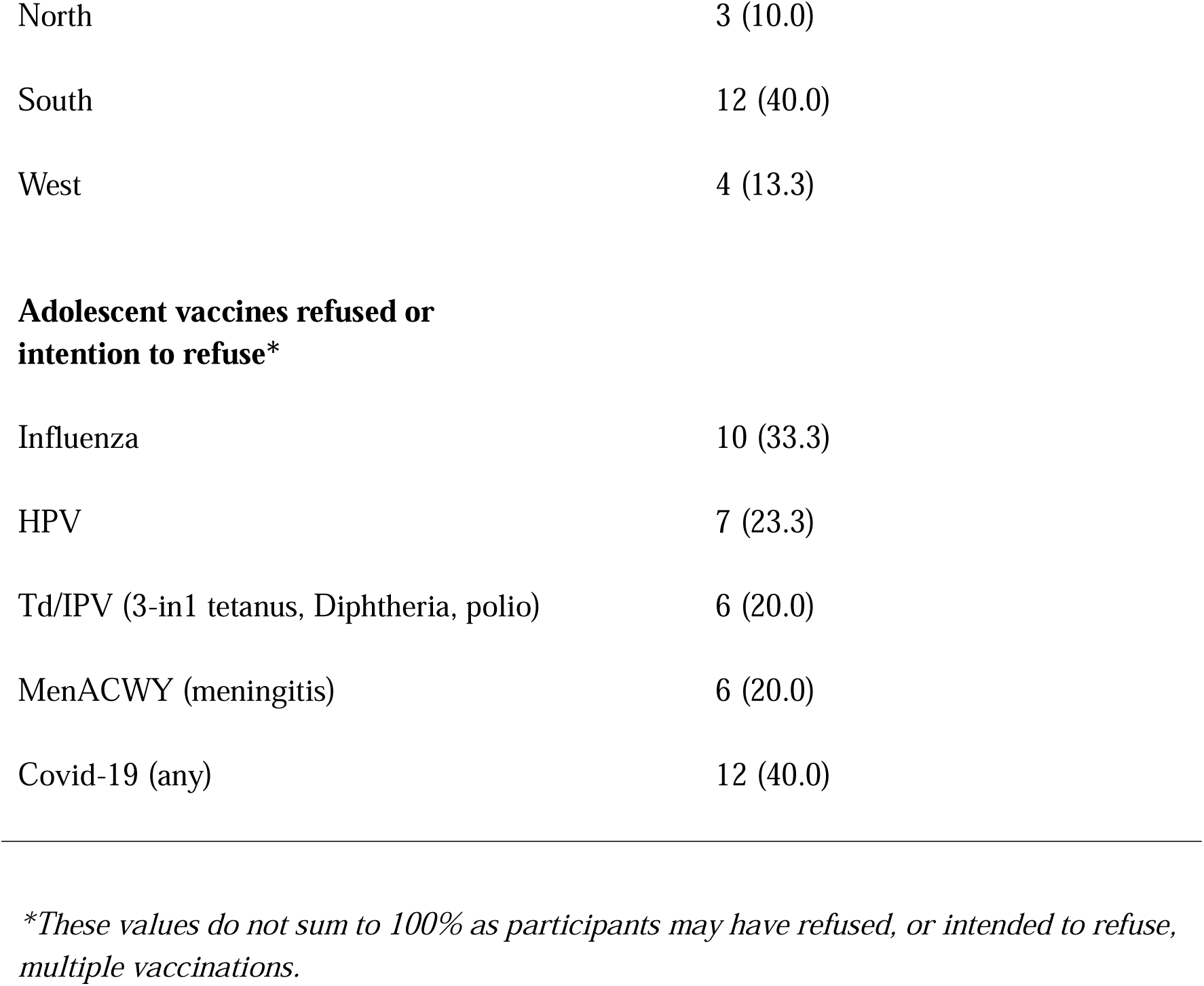
Participant characteristics and vaccine refusal

Following analysis of the data, five themes were identified (Figure 1):

1. Understanding vaccines
2. Fear and comfort
3. Autonomy and control
4. Information and rumours
5. Identity and social norms

**Figure 1:**
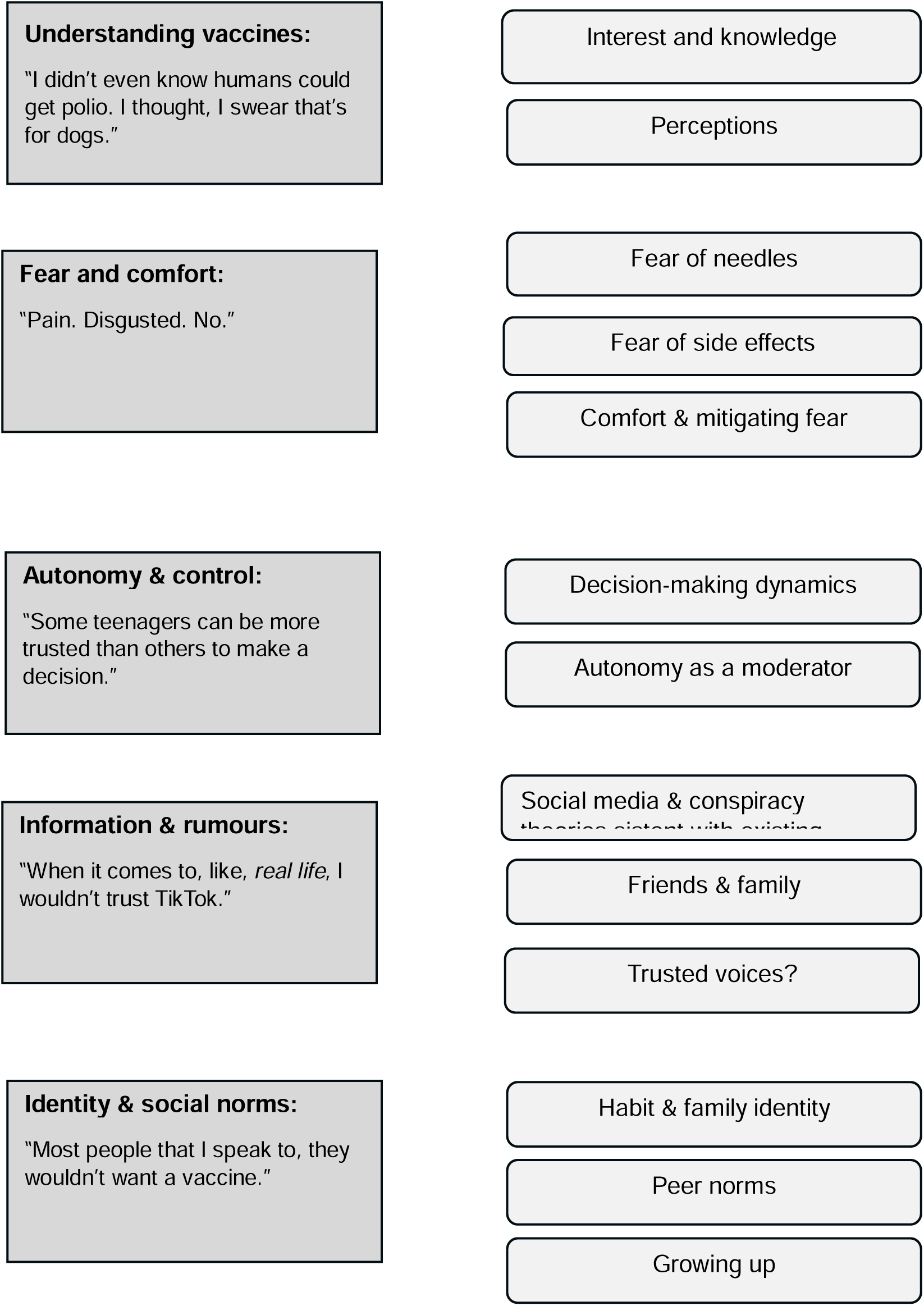
Summary of themes

### Themes

#### 1. Understanding vaccines: “I didn’t even know humans could get polio. I thought, I swear that’s for dogs.”

This theme captured adolescents’ level of interest in, and knowledge about, vaccines and vaccine preventable diseases (VPDs). Alongside knowledge, it described perceptions of vaccines and VPDs.

##### a) Interest and knowledge

For some participants, vaccines were not something they had considered, either individually, or through discussion with their peers:

> “*It’s not the fact that it’s not interesting… But I just think that [teenagers] … they’d rather talk about, uh, like, football, for example, or anything… Like, anything else*.” (P24)

Those who had an interest in science or had studied vaccines in more detail demonstrated enthusiasm for vaccines, e.g., “*It’s very interesting as a subject*” (P2). Participants generally had a good basic understanding of how vaccines worked. Participants who were vaccine acceptant often assumed their vaccine-hesitant peers simply lacked knowledge about vaccines, e.g., “*I think they need more knowledge on what the vaccine really is and what it does for them.”* (P6). However, vaccine hesitant participants also demonstrated good knowledge about vaccines:

> “*It’s building an immune system … Like I know what this vaccine is for. And I know why I chose not to take it.”* (P30)

Participants had little understanding of why routine vaccinations were targeted at their age group, and some VPDs were entirely unfamiliar to them, e.g., “*I didn’t even know humans could get polio. I thought, I swear that’s for dogs or something*.” (P16) Although they recognised Covid-19, influenza and cancer, not all participants knew what HPV was (“I think it might’ve been, like, HIV or something” (P1)) or that it was linked to some cancers. This was particularly clear amongst HPV vaccine-refusers. Overall, participants felt that current vaccine education in school was tokenistic, e.g. *“They will have like a five-minute talk to us … You’d have to look it up yourself, I guess”* (P21). Participants felt they should be able to learn about vaccines, VPDs and side effects from a younger age. Specifically, participants wanted the opportunity for discussion rather than didactic vaccine lessons and the opportunity to question trusted healthcare professionals in school. Several proposed an off-curriculum event:

> *“There should be a day … in school, where… someone comes in and they, like, talk to the classes and, like, people can ask questions … ‘Cause not everyone likes asking questions. So, if one person can ask, then they can all hear”* (P16, refused all vaccines)

##### b) Perceptions

In the absence of knowledge about VPDs, participants had developed their own perceptions of the diseases, sometimes based on ill-defined ‘feelings’ about them, e.g., “*I feel like tetanus is much more likely to be a danger to you*” (P3). Commonly, participants categorised vaccines as ‘serious’ and ‘non-serious’, based on the perceived severity of the disease. While for a minority Covid-19 was seen as deadly, e.g., *“COVID was way more serious. It was global.”* (P30), for others Covid-19 was inconsequential, e.g., *“The first time I got [Covid], I was ill for three hours, and then I was fine for the rest of the week.”* (P16). Where participants did not see COVID-19 as ‘serious’ for them, they nonetheless sometimes appreciated the severity for others who they wanted to protect. E.g. “*I wanted to protect my grandma and stuff like that*.” (P26). This concept was not mentioned by participants in relation to other vaccines. Yet the concept of herd immunity was not mentioned by any participants in relation to other vaccines. Influenza was also seen as less serious, e.g., “*Maybe it won’t be the end of the world if I get the flu …”* (P15), regardless of whether it was seen as safe, “*Flu vaccines, they’ve been researched a lot more, but I still just can’t see myself getting one, because I don’t feel the need to*” (P21). As well as not perceiving influenza as severe, some felt there was a lower likelihood of catching it, e.g. “*say if it was something like flu that you’re not a 100% going to get …”* (P8). Both influenza and Covid-19 (and the vaccines to protect against them) were downplayed with the use of minimising language, e.g., “*it was* just *the flu one*” (P9) “*[Covid is] like* just *a bad cold*” (P16), and, perceived as less serious, the vaccines were viewed as unnecessary, e.g.,

> *“There are some vaccines that you need to have, like… for your safety. But then maybe ones like, ones like the COVID vaccine and the flu vaccine, which probably aren’t that dangerous to you if you’re like my age …”* (P3)

Where participants associated vaccines with more serious diseases such as cancer, they were more motivated to accept the vaccine, e.g., “Obviously, I don’t want to, to get cancer, so I was, like, yeah, I will take the HPV vaccine.” (P21). Indeed, some participants saw themselves as lucky to have the opportunity to prevent some forms of cancer:

> *“Just knowing that if there is a way to prevent something like cancer, then I’m not going to pass up that opportunity, because not everyone gets that kind of opportunity.” (*P15)

#### 2. Fear and comfort: “Needle. Scared. Sports hall.”

This theme described emotional responses to vaccines, including fear of needles and of vaccine side effects. It also included the coping methods participants used to comfort themselves and others in the face of vaccine-related anxiety and the perceived absence of support from adults.

##### a) Fear of needles

Needle fear dominated the discourse for many participants. At the start of the interview, participants were asked to give three words related to vaccines. Needle was the most commonly shared word, mentioned by thirteen participants often in a negative context, e.g., “*Needle. Scared. Sports hall*” (P11). Participants used violent language to describe injections, e.g., “*stab*” (P5); “*being poked with a bit of metal”* (P15). With few exceptions, needles were associated with pain. However, needle fear existed on a spectrum, ranging from minor concern to needle and blood-related disgust, e.g., “Pain. Disgusted. No” (P19). For some, anticipating an injection induced anxiety, e.g., *“It just leaves a sort of feeling of dread in me for the days before it”* (P15). In some cases, this anxiety was unwarranted, and the experience was not as bad as had been anticipated. For others, especially where they experienced severe needle fear, repeated negative needle experiences perpetuated their anxiety and made them less likely to accept vaccines in the future:

“*It was really bad. I started crying … yeah, I think after that I don’t think I’d get one if I didn’t need to.”* (P27).

Other than brief reassurance from nurses, participants felt their needle fear was dismissed by adults, and those experiencing needle-related anxiety felt they were given little support from parents.

##### b) Fear of side effects

Side effect concerns were less pervasive than needle fear. For some, fear of side effects was emotion and observation-driven, especially where participants described a friend or relative who had experienced a vaccine-related death or severe injury. In such cases, vaccines were viewed negatively. However, other participants took a more measured approach to side effects, evaluating likelihood and severity to inform their decision. While some felt vaccines were worth the risk, others disagreed and refused vaccines due to side effect concerns, but in both cases, this was a reflective decision. Perceptions of what constituted a severe adverse effect were subjective, from “*being dizzy, sick*” (P18), “*losing a limb*” (P11); to “*spots and…death*” (P2). When asked where side effects come from, there was evidence of adolescents rationalising and reframing symptoms. While some attributed side effects to “*having a bad reaction to the germs”* (P8), others thought there could be psychological explanations such as anxiety, or expectation of side effects. One participant thought side effects could be explained by the misattribution of existing symptoms, and several participations made efforts to reframe side effects positively, as reassurance that the vaccine was working:

> *“Well, I mean, nobody wants to be sick, but I mean it’s good because your body is responding normally: you’re fighting it.”* (P30)

##### c) Comfort & mitigating fear

For adolescents, vaccines in school might be their first independent medical experience. In the absence of parents, friends, particularly females, often took on the role of comforters during vaccinations:

> *“[My friend] just came, and she sat next to me. And she held my hand. And she just talked to me”.* (P15)

Some participants felt more could be done to make vaccination comfortable for adolescents, including offering privacy or using diversions such as videos or music during school vaccination programmes.

#### 3. Autonomy and Control: “Some teenagers can be more trusted than others to make a decision.”

This theme incorporated agency and desire (or not) for control over vaccine decisions. It described different vaccine decision-making dynamics between parents and adolescents and shows how autonomy can both drive and disrupt vaccine intention and behaviour.

##### a) Decision-making dynamics

While some participants had full control over vaccine decisions, others described a more collaborative decision-making process, researching the pros and cons of vaccines with parents and making a joint decision. Other participants had no say at all, e.g. *“[My mum] just picked for me. I think she just picked no … I don’t really have a say.”* (P22). Some participants were indifferent to having agency on vaccines, especially if they were disinterested in the topic. Others expressed anger or frustration at being excluded from the decision-making process. Overall, most participants felt adolescents should at least have some say over vaccine decisions because it directly affected their own bodies and health, e.g. “*The idea of someone else saying … you can or can’t have it …is a bit, um, silly … it’s not them being infected, it’s you being infected.”* (P24) However, participants often felt the nature of the VPD should dictate the level of adolescent autonomy. Some felt they should be given more of a say where vaccines were seen as more serious:

> *“Participant: I might want to be, like, involved in, like, the big … vaccinations and stuff. Researcher: What do you think is a big vaccination? Participant: Like, a cancer one …”* (P12)

Others felt the opposite: that they were happy to decide about less serious vaccines but deferred to their parents on more ‘serious’ vaccines.

##### b) Autonomy

In this dataset, autonomy could both drive and disrupt vaccine uptake. Parental control over vaccine decisions often, but not always, determined uptake. Some vaccine-acceptant participants had vaccines refused on their behalf by vaccine-hesitant parents:

> *“I mean, I was relatively on board, but I think my parents were quite unsure, so I ended up not having it.”* (P7)

A minority recognised they had ultimate agency as they could override parental decisions in school, either to have the vaccine despite their parents’ wishes, or to reject the vaccine despite their parents’ consent e.g., *“I could just go to school and say I don’t want it. And then [my parents] can’t do anything”* (P15). However, this was rare: adolescents mostly viewed parents as having ultimate control because they signed the consent forms for vaccination.

#### 4. Information sources and rumours: “When it comes to, like, *real life*, I wouldn’t trust TikTok.”

This theme described information sources beyond formal education informing vaccine perspectives, including social media, misinformation and conspiracy theories. It described trusted (parents and the NHS) and less trusted (friends and the government) sources of information.

##### a) Social media & conspiracy theories

Though all participants used social media, they frequently denied seeing vaccine content online. They viewed social media as a source of entertainment, rather than information and would ‘scroll past’ topics relating to vaccination. Consequently, participants denied social media influenced adolescent vaccine attitudes. They knew social media was untrustworthy and distinguished between the online world and reality, e.g., “W*hen it comes to, like, real life, I wouldn’t trust TikTok”* (P5). Paradoxically, many assumed other adolescents were influenced by vaccine content on social media, e.g., *“TikTok is really, like, influential … they’re going to believe everything”* (P5). Often, the adolescents ‘othered’ their generation, describing them as “*a bit gullible on social media sometimes*” (P4) and the source of misinformation, e.g., “the current generation love to gossip …” (P16). Whilst most denied seeing vaccine rumours online, many were familiar with common Covid-19 vaccine conspiracy theories. Where participants were vaccine acceptant, rumours were viewed as comedic, and for vaccine-refusers, rumours reinforced their established belief that vaccines were harmful. However, those who were vaccine hesitant were more vulnerable to conspiracy theories. For them, rumours raised questions and introduced doubts:

> *“It made me think … maybe that is true. But at the same time, I feel like if … the government knows loads of people are dying … it would be more in their favour to find a solution”* (P23).

Worryingly, there was a suggestion from some that Covid-19 vaccine-rumours made adolescents less likely to accept future routine vaccinations, e.g., “*I think maybe people heard about these rumours about the Covid vaccine. And then they thought the same thing about like all the vaccines”* (P29).

##### b) Friends and family

While friends were not seen as a reliable source of information, parents generally were. Most argued that what their parents said had a far greater influence on their vaccination behaviours than what they saw on social media:

> *“I remember, like, scrolling on TikTok and everyone saying that the vaccines are fake … I was just like, okay, believe what you want to believe. I just, I just follow what my mum tells me to do [laughter].”* (P5)

With some exceptions, parents were by far the biggest influence on participants’ vaccine behaviours. However, not everyone was convinced that their families were knowledgeable e.g., *“My dad doesn’t know much about vaccines [laughing] … my sister’s a bit clueless as well.”* (P1), and some identified parents as the source of vaccine rumours. Some doubted their parents’ ability to spot vaccine misinformation:

“*Like parents … you know how you’re not meant to believe everything you see? Well, clearly not. Because they believed everything they saw. And they were so against the Covid vaccine.”* (P27)

##### c) Trusted voices?

Most saw the National Health Service (NHS) as a reliable and trusted source of information, with some giving examples of how they used the NHS website. Given this trust, participants wanted the opportunity to discuss vaccines with healthcare workers in school rather than being left to research on their own, e.g., “*I wouldn’t be going on my phone and finding out myself something that could not be true”* (P15). Information leaflets given to adolescents during vaccination were also viewed as reliable and sometimes useful. However, not all participants read the leaflet, either because it was not interesting to them or because it was emailed to their parents, and not to them. This was sometimes seen as problematic, denying adolescents the opportunity to make an informed choice themselves, e.g. *“[They] went in there with, like, nothing in their head about it … they just took whatever they were given.”* (P20)

While the NHS was trusted, attitudes towards the Government were more ambivalent. Some trusted the Government, but others were suspicious, e.g., “*“Sometimes I do feel like the Government can mislead people … they wanna have more control over people …”* (P23).

When asked what the NHS and Government could do to improve adolescent vaccine uptake, most wanted to be treated with respect, and for evidence to be transparent, e.g., “*Try and show the fact that you are telling the truth*” (P24). They rejected directives, arguing that with the appropriate evidence, adolescents could be trusted to make good decisions.

#### 5. Identity and social norms: “Most people that I speak to, they wouldn’t want a vaccine.”

This theme describes the role of social identity in vaccine choices. It captures the impact of social norms on vaccine attitudes and behaviours, including collective attitudes towards vaccines, and using peer group norms for self-reassurance. It also describes how maturity impacts vaccine attitudes.

##### a) Habit and family identity

Habit, or established behaviour, was a key heuristic in vaccine decisions:

> *“My friends, like, asked, are you getting the flu thing? And ‘cause I know I don’t normally, I just said no, and then I asked when I got home, and I wasn’t.”* (P16)

Often this was based on a family vaccine identity, which might be established by parents, or sometimes grandparents, e.g., “My grandpa does the same thing. He tells my mum … that they need to get their vaccines, and everyone gets their vaccines” (P5). Participants used first-person plural pronouns *(“we’re all pro-vaccine in this family”,* P2) to describe a collective attitude. Where participants disagreed with their parents on vaccines, they were less likely to use affiliative language, e.g., “*they’d probably say why I should have it and all the good things about it. And probably try to persuade me to get it.”* (P13). Participants who strongly identified with their family described mirroring parental vaccine behaviours during the Covid-19 pandemic, e.g., *“So, if they’re not going to have it, then maybe I shouldn’t either”* (P16). Where adults were not eligible for routine vaccines and so behaviour could not be copied, participants drew upon established family identity to shape their own decisions or to make sense of parental decisions.

##### a) Peer norms

While peers were not seen as a reliable source of information, peer norms were used to validate vaccine behaviours, e.g., “A *lot of my friends and classmates have been getting it. So, I was like, well, if they’ve decided to get it, then it should be fine”* (P9). Most participants’ vaccine choices were in line with their peers, whether vaccine acceptant or hesitant. Where participants found themselves misaligned with their peers, this made some rethink, e.g. “[*It*] *might make me like, reconsider my decision. Like maybe, maybe”* (P30). Others, particularly those disinterested in vaccines, felt whether their friends had vaccines was irrelevant. There were also examples of peer social contagion. Peers might exacerbate anxiety before vaccines, e.g., “*everyone else will be like, oh, God, are we all going to faint or something.”* (P11), or experience shared symptoms following a vaccine, e.g., *“Everyone felt like… they had a bit of a headache and stuff”* (P1). In one example, following a traumatic incident at school where an older pupil collapsed, peers reinforced assumptions that the vaccine was responsible:

> *“Everybody obviously thinks it was [the vaccine], because it was the same day, perfectly healthy person, and then just collapsed in the middle of a rugby match … Nobody in my year that I know has had [the vaccine].”* (P21)

##### b) Growing up

Adolescence is a transitional age, from childhood to adulthood. Younger participants in this dataset saw themselves as children, e.g., “*teenagers are more, like … we’re children, I guess. Adults are adults.*” (P1). By contrast, those in later adolescence often saw themselves as emerging adults. Some participants, predominantly females, were striving for independence and saw taking responsibility for health decisions as part of that journey. For some, views on vaccines formed part of their emerging adult identity, e.g., vaccines were *“just not something I would do”* (P22). Younger participants were more likely to be influenced by emotional, short-term responses to vaccines, e.g., needle pain, and equally celebrated short-term benefits, e.g., avoiding lessons “*if [the vaccine] happens to overlap with PE, because then I get to not have to run around in circles for ages”* (P2). Older participants seemed more able to rationalise their fears and self-motivate than younger participants, e.g., “*Sometimes [you] just have to suck it up, you know?* (P27). Younger participants struggled to imagine a future in which they would make vaccine choices independent of parents, e.g., *“I feel like when I’m older, my mum will still be telling me that I need to get my vaccines”* (P5), whereas older participants were more future-oriented, recognising the long-term benefits of vaccination. Older participants recognised that giving adolescents the skills to make informed vaccine decisions would not only promote individual health, but also support the well-being of future generations:

> *“[We need to] educate teenagers about vaccines and then they can take that with them throughout later life and then when they become parents and stuff.”* (P28)

## Discussion

This study explored barriers and facilitators to vaccination in adolescents. It supports prior research that needle fear and concern about side effects are barriers to vaccination for adolescents [13, 38, 39]. It offers novel insights into how identity formation, autonomy and social norms impact adolescent vaccine intention and behaviour. This study was designed, piloted and analysed in partnership with adolescents, allowing us to collect rich and detailed data, and to offer adolescent-informed interpretations of the results.

In this study, adolescent development and identity formation shaped vaccine behaviour. In line with broader literature on adolescent health [27], in younger participants we saw little focus on the future, and consequently poor perceptions of long-term risks from VPDs. Younger participants were more likely to act on emotional responses to vaccines, especially needle fear, and struggled to regulate these emotions. They relied on a family approach to vaccination with parents acting as a key source of information, making vaccine decisions or supporting collaborative decision-making. Younger participants based their vaccine attitudes on a group family approach to vaccination. On the other hand, older participants who were moving toward adulthood and independence were more able to envisage the long-term benefits of vaccines, not just for their own health but also their ability to make informed health decisions having a positive impact on their children and therefore future generations. While retaining their family identity toward vaccines, other influences had been incorporated (education, friends) to form an emerging adult attitude to vaccines which they felt was unlikely to change in the future. This shows how important it is to establish positive, well-informed attitudes toward vaccines from a young age.

Autonomy was both a driver and a disrupter of vaccine behaviour. As in broader literature [25, 40], in this study we found decision-making dynamics between parents and adolescents to be complex, ranging from complete autonomy to none. The adolescent-parent decision-making dynamic was influenced by the age and maturity of the child, their level of interest and knowledge about vaccines and a family approach to collaborative research and decision-making, and the willingness of the parent to cede or share decision-making responsibility. Despite evidence from wider literature that involving adolescents in vaccine decisions is associated with greater acceptance [25, 41], in this study there was no consistency between adolescent agency and vaccine uptake. Crucially, as well as driving vaccine uptake, autonomy also sometimes disrupted vaccine intentions (acceptant or otherwise). The ‘intention-behaviour gap’, that is the discrepancy between intention to do a healthy behaviour, and actually carrying out the behaviour [42] is widely described in health psychology [43, 44] . In adults, factors disrupting vaccination intention include forgetting, lack of time and emotional responses overriding planned behaviour [45]. This study shows autonomy (or lack of autonomy) can also disrupt vaccine intentions in adolescents. A parent might override an adolescent’s refusal to be vaccinated, especially if this refusal was based on an emotional response e.g., needle fear. However, parents could also override adolescent vaccine acceptance, with broader research suggesting adults may also refuse vaccines for their children based on cultural beliefs, fear of side effects and mistrust of authorities [11, 46, 47]. This highlights the need to inform and engage both parents and adolescents about forthcoming vaccines and vaccine decisions. In line with prior research [40] adolescents believed they should have some say in vaccine decisions as it was their health being directly affected. Given adolescents are typically inexperienced in healthcare decisions [27], involving adolescents in vaccine decisions would not only directly impact their health, but would also give them the skills to critically appraise health information and make informed independent decisions in the future.

Peers and social identity are known to be more influential in adolescence than at any other time in the life course [18]. Broader literature has demonstrated peer influence on health risk behaviours, e.g. substance use [48]. By contrast, we saw little evidence of peers directly impacting adolescent vaccine acceptance or rejection, presumably in part because most vaccine decisions were made at home. Peers were neither seen as a reliable source of information, nor as someone to copy or turn to for vaccine advice. Peers nevertheless played an indirect role in adolescent vaccination: although not directly copying their friends, peer norms offered an opportunity for validation of existing vaccine decisions. Moreover, there was evidence of social contagion, where attitudes and behaviours are transmitted from one individual to another [5]. Friends could sometimes exacerbate pre-vaccine anxiety or experience shared symptoms after vaccines. However, friends’ influence was also positive, taking on the role of carers in the absence of parents during in-school vaccines, offering comfort and reassurance. The complex roles of social identity, individual agency and social networks suggest that vaccine interventions involving family and friends could increase vaccine uptake [5].

Whilst it is easy to assume that better education on vaccines would increase vaccine uptake, in this study we found adolescents who were knowledgeable about vaccines could be both vaccine hesitant or acceptant. Most participants had a basic understanding of how vaccines work, but less knowledge about the diseases against which vaccines offered protection. In the face of these knowledge gaps, younger participants in particular assigned binary definitions to VPDs as serious or non-serious, and this dictated both whether to accept that vaccine, and who should make the decision. Both influenza and Covid-19 were seen as less serious diseases for adolescents, and therefore vaccines to protect against them were perceived as unnecessary. HPV, when associated with cancer, was defined as a serious vaccine which ought to be accepted. This maps to wider literature, which shows an association between increased HPV knowledge and positive attitudes toward [49], and increased uptake of [50], HPV vaccination. In our study, participants who had not accepted the HPV vaccine were often unaware that either it was more effective if given at a younger age [51], or that it protected them from some forms of cancer. Emphasising both may therefore increase uptake. Overall, current vaccine-related education in school was viewed as passive and tokenistic. As in similar studies [3, 20, 52], adolescents wanted the opportunity to discuss vaccines with healthcare experts from an earlier age.

Every participant in this dataset said they used at least one social media platform. This is not surprising, given 95% of 13–15-year-olds and 98% of 16–17-year-olds in the UK use social media [53]. As in other research [20], adolescents denied being influenced by vaccine content online, instead implicating adults as the ones who engaged with and shared vaccine misinformation. Other qualitative studies have shared similar findings, e.g., with adolescents acting as ‘debunkers’ for older relatives on Covid-19 vaccine misinformation [54] . However, with indirect questioning, e.g., ‘Why do you think *other* teenagers say no to vaccines?”, participants othered their generation, subscribing to age-related stereotypes and blaming social media. Perhaps adolescents believed admitting to being influenced by social media would lead to them being judged negatively by researchers and so projected those behaviours onto their peers. Since adolescence is such an intense period of social redefinition and cognitive development, adolescents in research can be sensitive to perceived judgment and display social desirability bias, reporting behaviours in a way that they perceive as socially acceptable [55, 56]. Even so, their arguments that vaccine information online was just not interesting were compelling, and adolescents gave multiple specific examples of alternative influences, supported by PPIE analysis.

Emotion-driven responses to vaccines such as fear of needles or of severe side effects were barriers to vaccination in this study, particularly for younger participants. This is unsurprising, given adolescents have less capacity than adults to override emotional responses [27]. Needle fear is a known barrier to adolescent vaccination [13, 57]. A recent meta-analysis reported needle fear prevalence in adolescents to be between 20 and 50% [58] and needle fear and pain were estimated to be a barrier to childhood vaccination in 5% to 13% of the general paediatric population and 8% to 28% the under-vaccinated paediatric population. In this study, needle fear ranged from low-level worry to extreme anxiety or disgust, and where participants reported negative needle experiences, they were less likely to accept future vaccinations. While needle fear does appear to decrease with age [58], it is distressing in the short term, and in the long term could lead to negative attitudes toward and non-compliance with future vaccinations and other medical procedures in adulthood [13].

Further, needle fear in parents can be transferred to future generations [59], perpetuating the problem. More could be done to improve the experience of vaccination for adolescents, including low-cost interventions to decrease anxiety such as distraction techniques, and support with understanding and managing emotional responses. In line with prior reviews [57, 58], we suggest more research into simple, scalable psychological interventions which may reduce needle fear in the adolescent population.

Concern about side effects was less dominant in this study than in the adult vaccine literature, but was still a barrier to vaccination for some, especially where this response was emotion-driven after witnessing other people experience severe vaccine injuries. Where adolescents considered side effect risks more reflectively, weighing up risks versus benefits, they were often vaccine acceptant. There was evidence of some adolescents attempting to rationalise symptoms and explore alternative explanations for side effects. This suggests that interventions which allow adolescents to explore side effects in more detail (balanced with an understanding of when to seek medical help) could be useful. In research with adults, interventions which supported participants to develop a more positive, adaptive mindset toward vaccines reduced stress and increased vaccination intention for unfamiliar vaccines [60, 61]. Our research suggests an opportunity to leverage and support adolescent interest in side effects. Discussing psychological explanations for needle pain or side effects, such as the nocebo effect, where individuals’ experience of side effects may be based on expectation, social observation or misattribution [16] could help adolescents to develop the skills to self-regulate their fears.

## Limitations

More than half of participants in this study lived in London or the southeast of England (n = 18). Furthermore, we struggled to recruit younger vaccine hesitant adolescents to this study and eventually recruited six participants via parent panel members of a market research agency. Some adults acting as gatekeepers for vaccine-hesitant adolescents refused to share details of this study. It is likely therefore that a portion of the current adolescent generation who are, or whose caregivers are, strongly anti-vaccine and anti-government, are not represented and therefore underserved by this research. Further, all participants in this study lived with at least one parent, and parents were a key influence on vaccine behaviour. More research is needed to understand how adolescents who live outside of a family unit (cared for adolescents for example) make effective vaccine choices.

## Conclusion

In conclusion, this co-produced study offers new insights into how adolescent vaccine attitudes develop during this unique period of intense physical and social change, distinct from childhood and adulthood. Overall, adolescent vaccine attitudes were complex and context dependent, with participants wanting more information on and involvement in vaccine decisions. This study suggests ways in which adolescents could be better supported to make informed decisions about vaccines, including improving knowledge of VPDs and supporting adolescents to manage emotional responses such as fear. The role of consent and autonomy were crucial, and encouraging the involvement of adolescents in vaccine decision-making could help adolescents to develop the critical thinking skills needed to make informed health decisions both now and in the future.

## Statement of authorship

**Angie Pitt:** Conceptualization; Methodology; Formal analysis; Data collection (interviewing); Writing - Original Draft; Funding acquisition.

**Professor Richard Amlôt:** Conceptualisation; Supervision; Writing - Review & Editing.

**Dr Catherine Heffernan:** Conceptualisation; Supervision; Writing - Review & Editing.

**Professor G. James Rubin:** Conceptualisation; Supervision; Writing - Review & Editing; Funding acquisition.

**Dr Louise E. Smith:** Conceptualisation; Methodology; Formal analysis; Supervision; Writing - Review & Editing; Funding acquisition.

## Ethical statement

The PNM Research Ethics Panel at King’s College approved this study (LRS/DP-23/24-39491). All participants aged 16 to 17 years gave informed consent. Where participants were aged 12 to 15 years, parents gave informed consent and participants gave informed assent.

## Data Availability

All data produced in the present study are available upon reasonable request to the authors.

